## Supplementary material for "Waiting times, patient flow, and occupancy density in South African primary health care clinics: implications for infection prevention and control"

### Appendix 1. Literature review to inform choice of method

#### 1.1. Methods

##### 1.1.1. Choice of data collection method

A literature review was undertaken to find a data collection method that:

1. Allowed collection of data on the movement of all visitors to the clinic, including those accompanying patients and children, and all clinic staff;
2. Allowed collection of data on movement in all parts of the clinic, including waiting areas;
3. Allowed collection of anonymised data about basic demographics of those attending (age group, sex, reason for attendance) and allowed for identification of those attending with a baby;
4. Did not involve the installation of additional technical (or other) equipment or infrastructure in clinics;
5. Could easily be moved between clinics;
6. Did not violate the privacy or confidentiality of clinic attendees or staff;
7. Was not excessively expensive; and
8. Ideally, could be used (after modification, if needed) in routine practice after the conclusion of the study.

##### 1.1.2. Searching, sifting, and inclusion and exclusion criteria

MEDLINE (via PubMed), Scopus, Web of Science, CINAHL, and Google Scholar, were searched using variations (including Medical Subject Heading [MeSH] terms) and combinations of the following terms: “queue”, “patient flow”, “waiting times”, “measurement”, “modelling”, “lean”, and “six sigma”. Results from the different databases were compared and duplicate records removed. Titles and abstracts were then hand-sifted; studies were included that described measurement of the physical movement of individuals through a defined structure or system (including, but not limited to emergency departments, hospitals, airports, and retail outlets); reported on the measurement of the movement of other objects through a defined structure or system (including reports of manufacturing processes and applications of traffic flow theory); attempted to measure the size and/or density of crowds of people, either indoors or outdoors; and reported on methods used to analyse similar data (including applications of queue theory). Articles were excluded that described the application of queue theory to the design or implementation of computing networks or systems (for example, studies of methods to improve management of web traffic) or that were purely theoretical (i.e., that did not describe the collection or analysis of data).

##### 1.1.3. Data extraction

Data were extracted by one individual; for articles reporting on data collection, information captured include the study setting, the objectives of the data collection exercise, the outcomes of interest, and a detailed description of the methods used to collect data. For studies reporting on analyses of previously collected data, information captured included any available details of how the data were collected, the outcomes of interest, a description of the analysis process, and any theory used to inform it. A pragmatic, snowballing approach was adopted: where an article described a method that was the same as or very similar to one that had already been documented, only major differences in setting and methods were recorded, instead of all the details described above. As this was a review of methods, findings from included studies were not extracted.

Records were organised using Mendeley and Microsoft Excel; Microsoft Access was used for record sifting and data extraction.

#### 1.2. Results

##### 1.2.1. Methods considered; criteria used to make choice

Supplementary table 1 summarises some of the methods that have been used to measure waiting times and patient flow in health care settings and Supplementary table 2 provides a summary of methods identified and the dimensions they could be used to measure.

**Supplementary table 1. Overview of methods used in previous studies (assigned to broad categories)**

| # | Category | Description/elements | Key references/examples |
|---|---|---|---|
| 1 | **Paper-based systems** | 1. Log times of patient entry and exit from the clinic<br>2. Simple, often paper-based system for tracking patients' movement around clinics (allows for estimation of time at a particular station as well as time spent between stations/in waiting rooms)<br>3. Observation of patient flow by research team<br>4. Direct observation/measurement of consultation times with various groups of HCW | Bachman 1997[1]<br>Ideal clinic[2]<br>Reagon 2010[3] |
| 2 | **Real-time location systems** | e.g., Radiofrequency identification systems used to track waiting times/patient movement within a facility | Singman 2015[4] |
| 3 | **Proximity sensors** | 'Protractor' – uses infra-red light to measure distance and relative body orientation of interacting users | Montanari 2018[5] |
| | | Social fMRI - includes a mobile (android-based) programme that records proximity to other phones | Aharony 2011[6] |
| | | iEpi – an application that collects data on social interactions (but only between people that have the application) | Aiello 2016[7] |
| | | Other, low-tech ways to estimate proximity/interaction (e.g., interviews, observation, diaries, etc.) | |
| 4 | **Systems for estimating queue sizes & durations** | Used widely in commercial settings: airports, retail; often involve infra-red technology/use of other sensors; use of proprietary software | https://www.airport-suppliers.com/supplier/qmetrix-gmbh/ |
| 5 | **Camera-based systems (video or still images)** | Used in retail; use 'video management system' as a baseline – can then provide information on ingress, egress, queue length, waiting time<br>Various levels of sophistication – can use machine learning/AI, face recognition; needs fairly heavy-duty computing infrastructure (1TB HDD server | www.allgovision.com |
| | | **Data-driven crowd analysis** – software that analyses video images of crowds and 'learns' patterns by performing long-term analysis in an off-line manner.<br>"…learn crowd motion patterns by performing long-term analysis in an off-line manner. The learned motion patterns can be used in a range of application domains such as crowd event detection or anomalous behaviour recognition. …The idea is that any given crowd video can be thought of as being a mixture of previously observed videos." | Rodriguez 2011[8] |
| | | **Density-aware tracking**<br>"Detecting and tracking people in crowded scenes is a crucial component for a wide range of applications including surveillance, group behaviour modelling and crowd disaster prevention. The reliable person | Rodriguez 2011[9] |

| # | Category | Description/elements | Key references/examples |
|---|---|---|---|
| | | detection and tracking in crowds, however, is a highly challenging task due to heavy occlusions, view variations and varying density of people as well as the ambiguous appearance of body parts, e.g. the head of one person could be similar to a shoulder of a near-by person. High-density crowds present particular challenges due to the difficulty of isolating individual people with standard low-level methods of background subtraction and motion segmentation typically applied in low-density surveillance scenes." | |
| | | **Structured vs. unstructured crowd tracking**<br>"In an unstructured crowded scene, the motion of the crowd appears to be random, with different participants moving in different directions at different times. That is, in such scenes each spatial location supports more than one, or multi-modal, crowd behaviour. For instance, a video of people walking on a zebra-crossing in opposite directions is an example of an unstructured crowded scene because, broadly speaking, at any point on the zebra crossing the probability of observing a person moving from left to right is as likely as observing a person walking from right to left. Other examples of such scenes include exhibitions, crowds in a sporting event, railway stations, airports, and motion of biological cells." | Rodriguez 2009[10] |
| **6** | **Measurement of structured or linear flow (e.g., traffic)** | **Measurements at a point or between two points**<br>• "This method is easily capable of providing volume counts and therefore flow rates directly, and with care can also provide time headways" | Traffic flow theory[11] |
| | | **Measurements over a short section**<br>• "All of these presence detectors continue to provide direct measurement of volume and of time headways, as well as of speed when pairs of them are used" | |
| | | **Measurements using (a) moving observer/s**<br>• "While the intention in this method is that the floating car behaves as an average vehicle within the traffic stream, the method cannot give precise average speed data. It is, however, effective for obtaining qualitative information about freeway operations without the need for elaborate equipment or procedures." | |
| | | **Measurements across a system/wide area**<br>• The limitation to all three systems is that they can realistically be expected to provide information only on speeds. It is not generally possible for one moving vehicle to be able to identify flow rates or densities in any meaningful way." | |
| **7** | **Broader/ systemic/ approaches to flow/ queue management** | **Queue science**<br>• "Patient flow in hospitals can be naturally modelled as a queueing network, where patients are the customers, and medical staff, beds and equipment are the servers.") | Armony 2015[12] |

| # | Category | Description/elements | Key references/examples |
|---|---|---|---|
| | | **Operations research/management**<br>• "Having time varying arrivals and heterogeneous patients that need to be treated in consecutive processing steps by several doctors, nurses and other employees, it is a complex environment to control."<br>• "Measurement of crowding: A fundamental weakness is the lack of a measurement gold standard. There is a weak literature base describing scoring systems of crowding." | Carmen 2014[13]<br>Higginson 2012[14] |
| | | **Various QI approaches**<br>• Linear process evaluation<br>• Action planning<br>• Patient flow simulator | Bean 2019;[15] Patient flow simulator: https://khp-informatics.github.io/patient-flow-simulator/ |
| | | **Game theoretical approaches**<br>• Patient satisfaction | McAdams 2014[16] |

**Supplementary table 2. Overlap between outcomes of interest and methods of measurement identified through literature review**

| **Outcomes** | **Numbers of people** | | **Time** | | | | **Proximity/density** | | | **Movement** | |
|---|---|---|---|---|---|---|---|---|---|---|---|
| **Method of measurement** | Numbers in and out of the clinic | Numbers in and out of different parts of the clinic | 'Average' time spent by individuals in the facility | Time spent by specific individuals in the facility | Time spent by specific individuals in different parts of the clinic | Time spent in proximity with other people/ other specific people | How/ how much people interact with each other | How closely people are grouped | Coughing/ other proxies for transmission | Average speed between key points/ through a particular area | Overall efficiency of the clinic |
| Motion sensors/footfall cameras/counting numbers in & out | **X** | **X** | **X** | | | | | | | **(X)** | |
| **Waiting/working time survey (paper-based or real-time location system)** | **X** | **X** | **X** | **X** | **X** | **(X)** | | | **(X)** | **X** | **(X)** |
| Proximity sensors | | | | | | | **X** | **X** | **(X)** | | |
| Cough sensors | | | | | | | | | **X** | | |
| Systems for estimating queue lengths (numeric size and duration) | | | | | **X** | | | | | **X** | **(X)** |
| Camera-based systems | **X** | **X** | **X** | **(X)** | **(X)** | | **X** | **X** | **(X)** | | |
| Measurement of structured or linear flow (e.g., traffic) | | | **X** | | | | | | | **X** | **(X)** |
| Queue science approaches | **(X)** | **(X)** | | | | | | | | **X** | **(X)** |
| Operations research/management approaches | **(X)** | | **(X)** | | | | | | | | **X** |
| QI approaches | **(X)** | | **(X)** | | | | **(X)** | | | | **X** |

**x** = reliably estimated; **(x)** = partially estimated

### Appendix 2. Additional methods

#### 2.1. Data collection

**Supplementary figure 1. Flow diagram illustrating data collection process.**

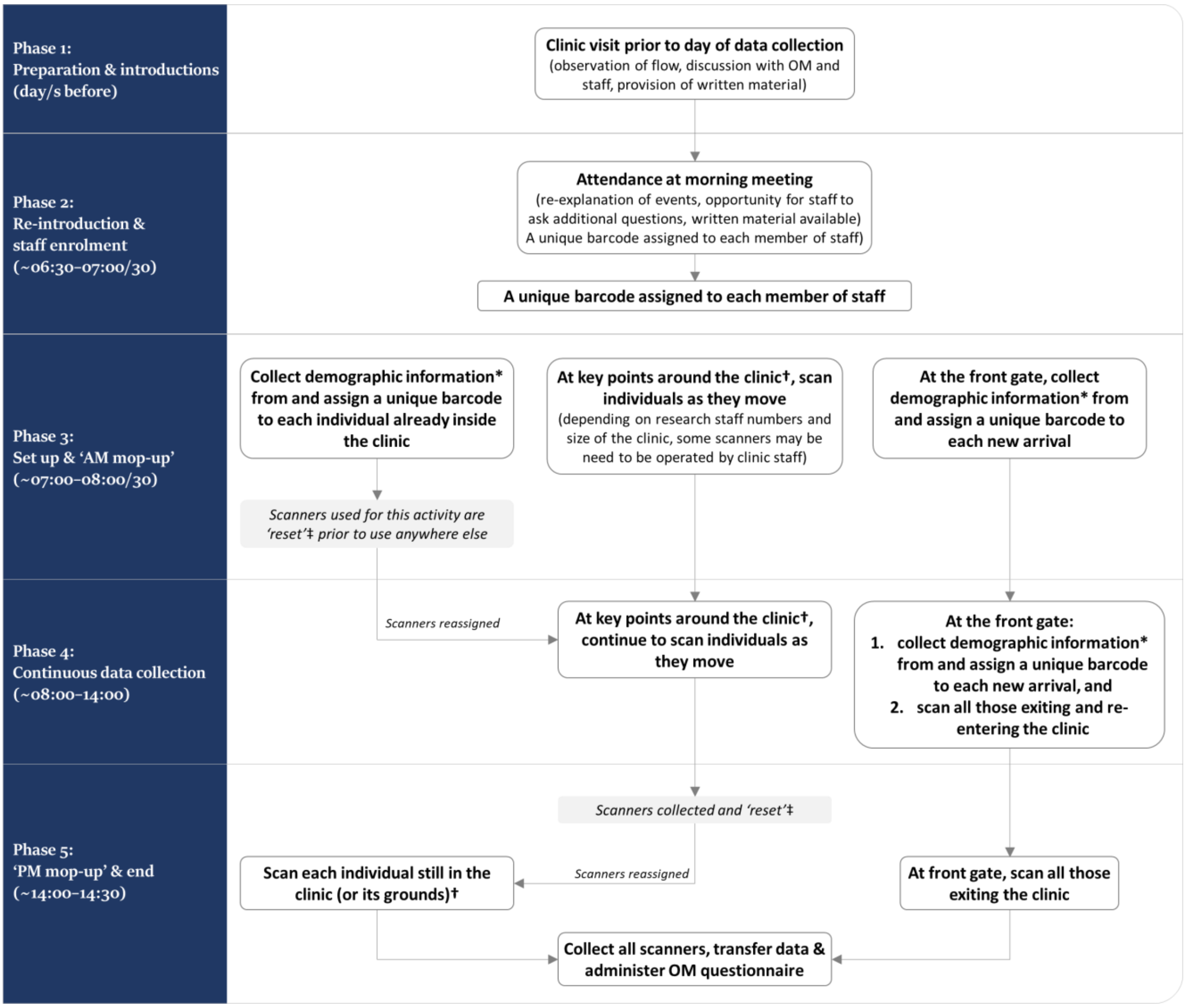

*Only non-identifiable information collected; participants were free to decline to participate
†Scanners were numbered and were 'location specific'. A record was kept of which scanner was used where.
‡A designated barcode was scanned three times in quick succession to denote reassignment of scanner. Time of reset was documented as well as original and new locations of the scanner.

#### Supplementary figure 2. Cards used for data collection from patients and clinic visitors (left) and from clinic staff (right)

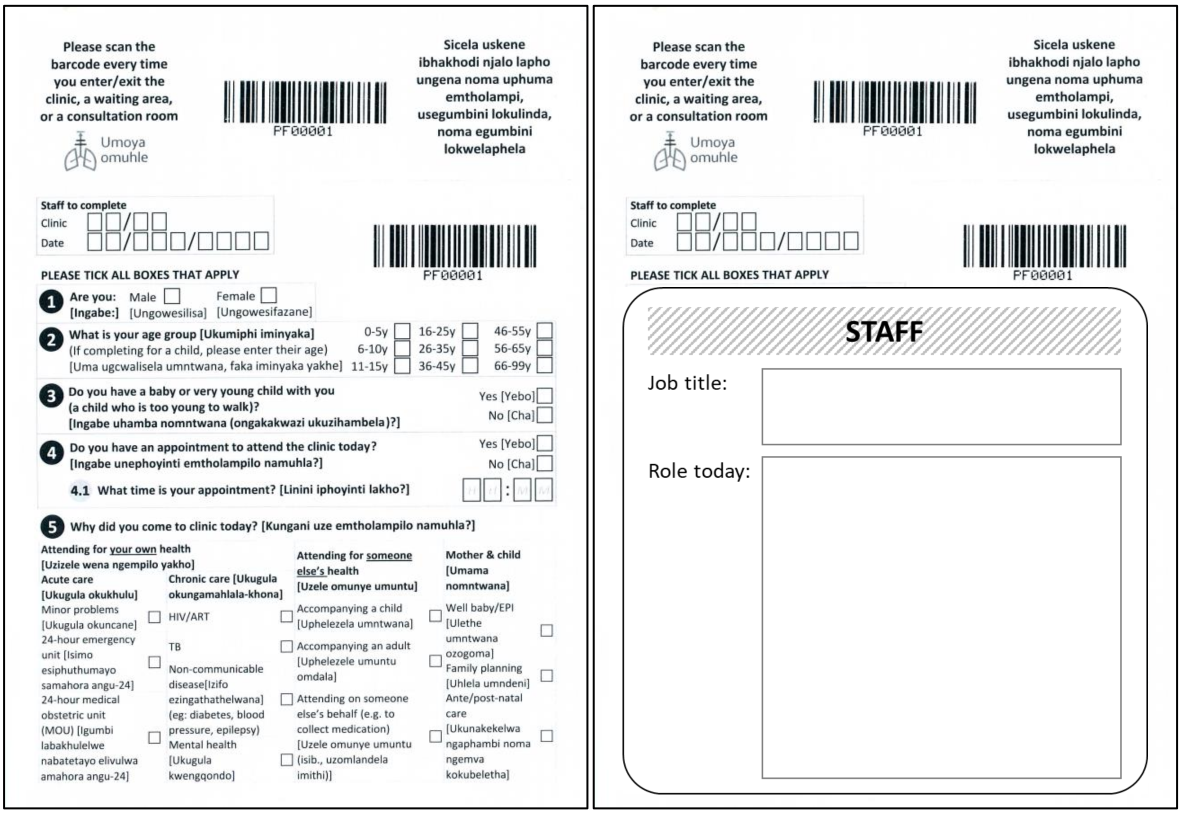

#### Supplementary figure 3. Schematic of an imagined small clinic, showing positioning of barcode scanners held by researchers and clinic staff

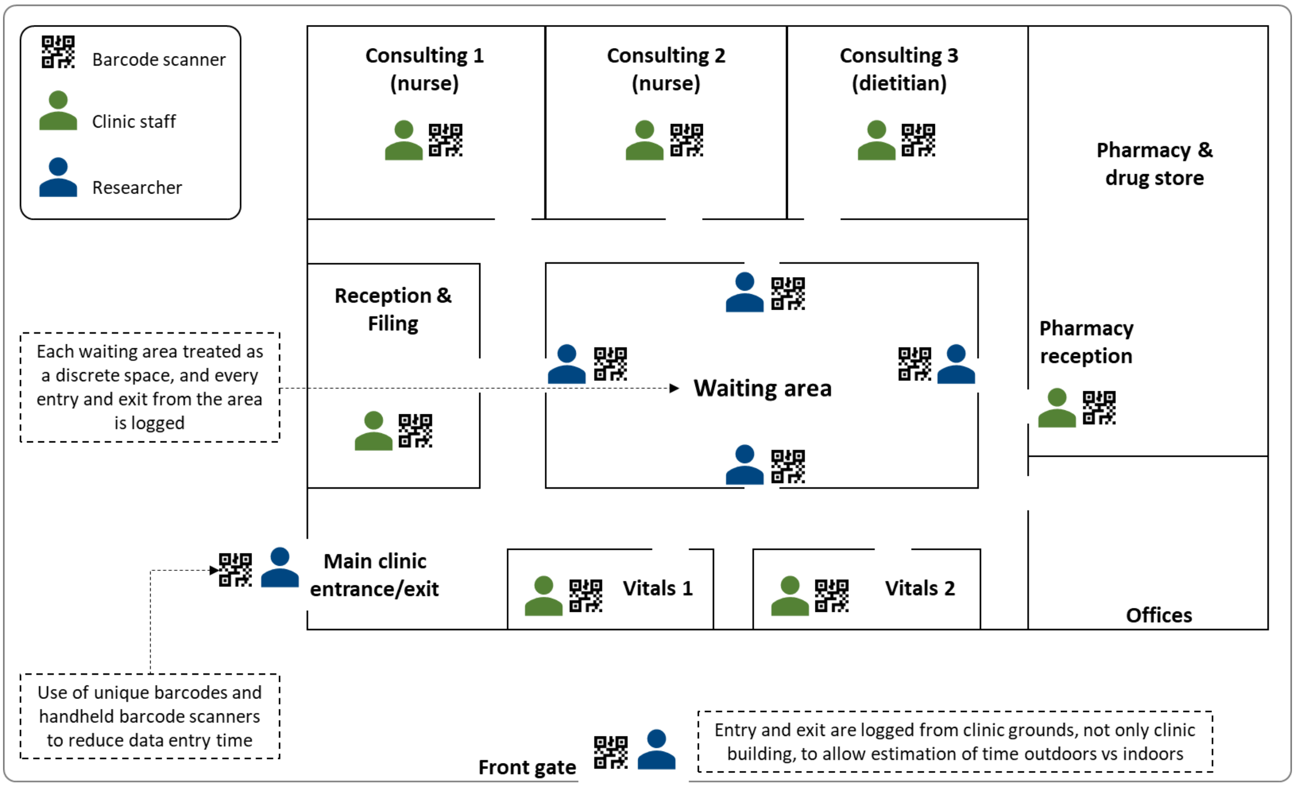

#### 2.2. Analysis

**Supplementary table 3. Number of clinics, number of individuals, and type of data used for each stage of the analysis**

| Analysis | Data used | Number of clinics | Clinics included | Number of data collection exercises | Additional exclusion criteria | Individuals included in analysis |
|---|---|---|---|---|---|---|
| Demographics | Original | 11 | KZN1, KZN2, KZN3, KZN4, KZN5, KZN6, WC1, WC2, WC3, WC4, WC5, WC6 | 12* | None | 2,903 |
| Time spent in clinic† | Imputed | 10 | KZN1, KZN2, KZN3, KZN5, KZN6, WC1, WC2, WC3, WC4, WC5, WC6 | 11* | None | 2,643 |
| Proportion of time spent indoors vs. outdoors‡ | Original | 9 | KZN1, KZN2, KZN3, KZN6, WC1, WC2, WC3, WC4, WC5, WC6 | 10* | Total visit time <5 minutes | 2,190 |
| Occupancy density of indoor spaces§ | Original | 3 | KZN2, KZN6, WC1 | 3 | None | 847 |

*Clinic KZN1 visited twice.
†Clinic KZN4 excluded as entry and exit from clinic not captured.
‡Clinics KZN4 and KZN5 excluded as coverage of all gates and indoor/outdoor doors not achieved.
§Suitable data only available from three clinics with multiple indoor waiting areas.
KZN: KwaZulu-Natal; WC: Western Cape

##### 2.2.1. Multiple imputation

Four key times were identified in the pathways that each clinic attendee took through the clinic.

1. **Time of arrival**

   The time that they first arrived at the clinic. This was assumed to be the time that their barcode was first scanned, for attendees who arrived after the start of data collection. The arrival time was set to missing if the attendee was already present in the clinic before the start of data collection, or if the first time their barcode was scanned was not at a clinic entrance (an external door or compound gate).

2. **Time at files**

   The time that the attendee obtained their patient file from the clinic reception desk. This was assumed to be the time that their barcode was first scanned at files, provided that it occurred before the first time that they were scanned at vitals or at a consultation room. The time was set to missing if they never scanned at files, or if they scanned at vitals or a consultation room before first scanning at files.

3. **Time at vitals**

   The time that the attendee has their blood pressure, heart rate, and respiratory rate measured. This was assumed to be the time that their barcode was first scanned at vitals, provided that it occurred before the first time that they were scanned at a consultation room. The time was set to missing if they never scanned at vitals, or if they scanned at a consultation room before first scanning at vitals.

4. **Time of departure**

   The time that the attendee left the clinic. This was assumed to have occurred at the final time that they scanned their barcode, provided it occurred at a clinic exit point (an external door or compound gate). The leaving time was set to missing for attendees who were still at the clinic at the end of data collection, or if their barcode was never scanned at an exit point.

In a small number of cases, times at files and/or vitals were missing not because the attendee did not scan their barcode, but because the attendee did not complete that stage. For instance, some attendees who were at the clinic to collect medicine only may have skipped one or both stages. In many clinics, patients on TB treatment also skip the files and vitals stages. In all ten clinics however, the majority of patients were required to pass through both files and vitals, regardless of their visit reason.

Missing times (Supplementary table 4) were imputed as interval-censored values, with lower and upper bounds of when the event would have occurred, using a sequential approach. First, arrival times at the clinic were multiply-imputed using 20 imputations. For attendees who arrived before the start of data collection, the lower and upper limits of the time of arrival were set to the clinic opening time and the start of data collection, respectively. For those who arrived after the start, the lower limit was set as the start of data collection, and the upper limit was the time that the attendee was first scanned. Second, the time at files was imputed, using the imputed arrival time as the lower bound of the interval and time at vitals (if observed) as the upper limit. If time at vitals was not observed, the upper bound was set to the earliest of the maximum time from arrival to files observed in that clinic, the time of leaving (if observed), the end of data collection (if not there at end) or, close of clinic (if there at end). Third, the time at vitals was imputed, using the imputed time at files as the lower bound of the interval, and the setting the upper bound to the earliest of the maximum time from files to vitals observed in that clinic, the time of leaving (if observed), the end of data collection (if not there at end), or the close of clinic (if there at end). Finally, the time of leaving the clinic was imputed, using the imputed time at vitals as the lower bound, and setting the upper bound to the earliest of the maximum time from vitals to leaving observed in that clinic, end of data collection (if not there at end) or close of clinic (if there at end).

**Supplementary table 4. Number of attendees and number with data missing for time of arrival, files, vitals, and departure at each clinic from which data underwent multiple imputation (n = 2,634)**

| Clinic code | Attendees, n | Arrival, n (%/attendees) | | | Files, n (%/attendees) | | Vitals, n (%/attendees) | | Departure, n (%/attendees) | | |
|---|---|---|---|---|---|---|---|---|---|---|---|
| | | Known | Missing (arrived early)‡ | Missing (other) | Known | Missing | Known | Missing | Known | Missing (left late)§ | Missing (other) |
| KZN1 (#1)* | 234 | 77 (33) | 10 (4) | 147 (63) | 87 (37) | 147 (63) | 61 (26) | 173 (74) | 91 (39) | 70 (30) | 73 (31) |
| KZN1 (#2)† | 417 | 264 (63) | 135 (32) | 18 (4) | 146 (35) | 271 (65) | 155 (37) | 262 (63) | 248 (59) | 129 (31) | 40 (10) |
| KZN2 | 170 | 130 (76) | 36 (21) | 4 (2) | 64 (38) | 106 (62) | 81 (48) | 89 (52) | 120 (71) | 47 (28) | 3 (2) |
| KZN3 | 270 | 183 (68) | 78 (29) | 9 (3) | 10 (4) | 260 (96) | 77 (29) | 193 (71) | 220 (81) | 34 (13) | 16 (6) |
| KZN5 | 347 | 241 (69) | 84 (24) | 22 (6) | 18 (5) | 329 (95) | 38 (11) | 309 (89) | 246 (71) | 89 (26) | 12 (3) |
| KZN6 | 218 | 128 (59) | 63 (29) | 27 (12) | 109 (50) | 109 (50) | 120 (55) | 98 (45) | 174 (80) | 34 (16) | 10 (5) |
| WC1 | 337 | 224 (66) | 65 (19) | 48 (14) | 0 | 337 (100) | 133 (39) | 204 (61) | 194 (58) | 79 (23) | 64 (19) |
| WC2 | 69 | 65 (94) | 2 (3) | 2 (3) | 52 (75) | 17 (25) | 31 (45) | 38 (55) | 56 (81) | 11 (16) | 2 (3) |
| WC3 | 120 | 56 (47) | 44 (37) | 20 (17) | 51 (43) | 69 (58) | 41 (34) | 79 (66) | 54 (45) | 38 (32) | 28 (23) |
| WC5 | 308 | 110 (36) | 158 (51) | 40 (13) | 33 (11) | 275 (89) | 27 (9) | 281 (91) | 176 (57) | 43 (14) | 89 (29) |
| WC6 | 144 | 93 (65) | 40 (28) | 11 (8) | 54 (38) | 90 (63) | 70 (49) | 74 (51) | 121 (84) | 17 (12) | 6 (4) |
| **Total** | **2,634** | **1,571 (60)** | **715 (27)** | **348 (13)** | **624 (24)** | **2,010 (76)** | **834 (32)** | **1,800 (68)** | **1,700 (65)** | **591 (22)** | **343 (13)** |

*First data collection exercise
†Second data collection exercise
‡Arrived before the start of data collection.
§Left after the end of data collection

Age, sex, clinic, reason for visit, whether there at start/end, and whether the attendee was first scanned in the morning (before 10am) were included in the imputation model. Twenty imputed datasets were created.

### Appendix 3. Additional results

#### 3.1. Demographics

**Supplementary table 5. Characteristics of primary health care clinics at which patient flow data collection exercises were conducted (n = 12)**

| Clinic code | Decade built | Location | PHC clinic or CHC | Estimated monthly head count, thousands | Date-time appointment system?* | Covered outdoor waiting area used as part of patient pathway? | Data collection exercises, n | Number of attendees included per exercise |
|---|---|---|---|---|---|---|---|---|
| KZN1 | 1990s | Semi-rural | PHC | 11–14 | No | Yes† | 2 | Exercise 1: 234; exercise 2: 417 |
| KZN2 | 1980s | Rural | PHC | 5–8 | No | Yes | 1 | 170 |
| KZN3 | 2000s | Peri-urban | CHC | 27–30 | No | Yes | 1 | 270 |
| KZN4 | 1980s | Urban | CHC | 27–30 | No | Yes | 1 | 269§ |
| KZN5 | 1980s | Urban | PHC | 4–7 | No | Yes | 1 | 347 |
| KZN6 | 2000s | Rural | PHC | 3–6 | No | No‡ | 1 | 218 |
| WC1 | 2010s | Peri-urban | PHC | 25–28 | Yes | No | 1 | 337 |
| WC2 | 2000s | Urban | PHC | 1–3 | Yes | No | 1 | 69 |
| WC3 | 1980s | Peri-urban | PHC | 2–5 | Yes | No | 1 | 120 |
| WC5 | 2010s | Urban | CHC | 27–30 | Yes | No‡ | 1 | 308 |
| WC6‖ | 2000s | Peri-urban | PHC | 25–28 | Yes | No | 1 | 144 |

*Often only for patients attending for selected services
†Used only for selected patients (‘chronic stream’)
‡Outdoor area not part of normal patient pathway; used primarily before the clinic opens
§Data collected from HIV/chronic unit only
‖Not officially a CHC but as large as and with an equivalent patient load to most CHCs.
CHC: community health centre; PHC: primary health care; KZN: KwaZulu-Natal; WC: Western Cape

#### 3.2. Total time spent in clinic

##### 3.2.1. Time of arrival

Most individuals arrived early: overall median time of arrival was 09:00 (IQR 07:53–10:27. This was similar across all clinics (range 07:36–09:36) and between provinces (Supplementary figure 4).

**Supplementary figure 4. Histograms showing distribution of imputed time of arrival at clinic by province (n = 2643; 20 imputations)**

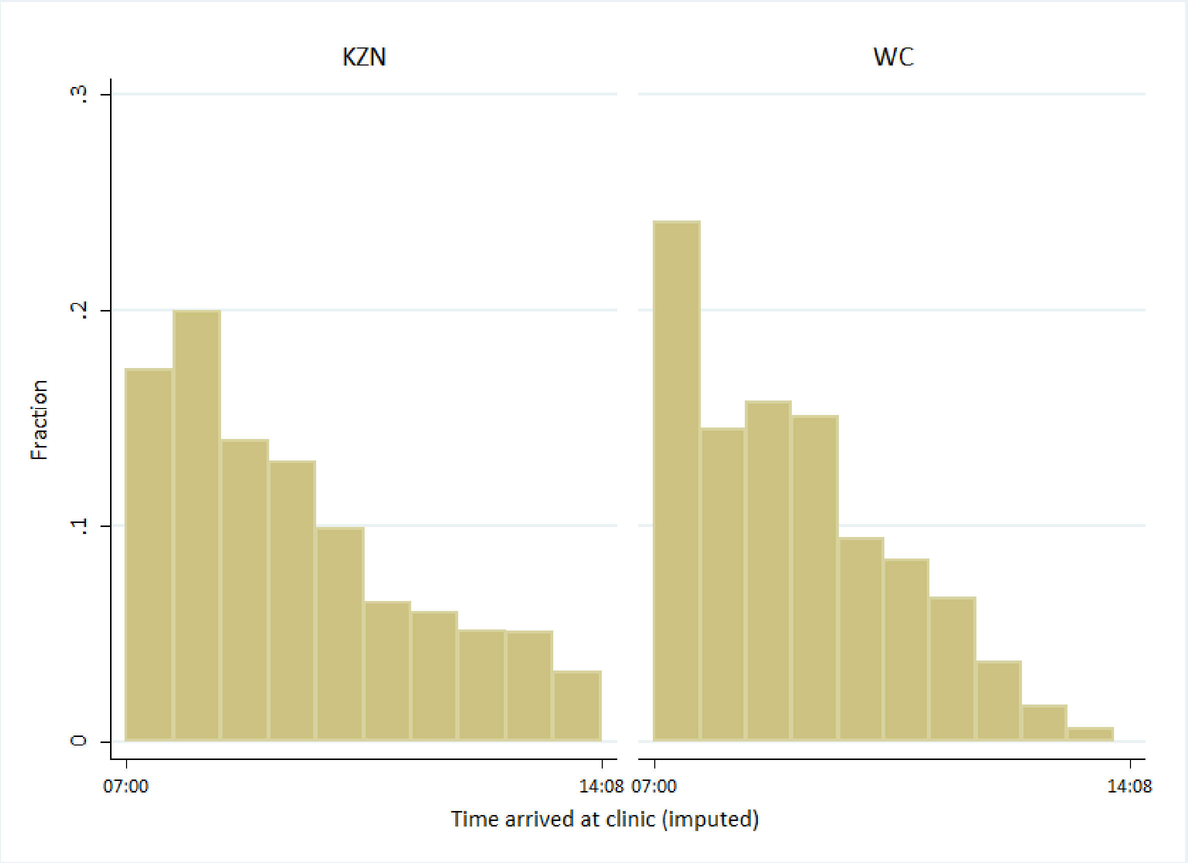

The shape of the relationship between time of arrival and time spent in clinic was examined using fractional polynomials, after extraction of one imputed dataset. The optimal second degree fractional polynomial of arrival time had the terms arrival time$^{(-2, 3)}$. However, there was no evidence that the optimal second degree or first degree fractional polynomial fit the data better than the linear model (p = 0.646 and p = 0.436, respectively) and time of arrival was included into the final model as a linear term.

**Supplementary table 6. Total time spent in clinic, by province and demographics (generated using imputed data; n = 2,634)**

| Demographic | Overall (n = 2,634) | | KwaZulu-Natal (n = 1,656) | | Western Cape (n = 978) | |
|---|---|---|---|---|---|---|
| | n (%) | Median (IQR) [range], HH:MM | n (%) | Median (IQR) [range] , HH:MM | n (%) | Median (IQR) [range], HH:MM |
| **Overall** | 2,634 (100) | 02:36 (01:36–03:43) [00:06–10:01] | 1,656 (100) | 02:33 (01:35–03:40) [00:09–10:01] | 978 (100) | 02:42 (01:37–03:49) [00:06–08:09] |
| **Sex** | | | | | | |
| Male | 783 (29.7) | 02:19 (01:25–03:32) [00:09–09:44] | 467 (28.2) | 02:16 (01:27–03:24) [00:11–09:44] | 316 (32.3) | 02:26 (01:21–03:39) [00:09–08:09] |
| Female | 1,851 (70.3) | 02:42 (01:41–03:47) [00:06–10:01] | 1,189 (71.8) | 02:38 (01:38–03:44) [00:09–10:01] | 662 (67.7) | 02:48 (01:46–03:55) [00:06–08:00] |
| **Age group** | | | | | | |
| <16 years | 381 (14.5) | 02:50 (01:45–03:48) [00:09–10:01] | 227 (13.7) | 02:45 (01:42–03:46) [00:09–10:01] | 154 (15.7) | 02:55 (01:51–03:48) [00:10–06:44] |
| 16–45 years | 1,703 (64.7) | 02:34 (01:36–03:43) [00:09–08:53] | 1,113 (67.2) | 02:30 (01:34–03:37) [00:10–08:53] | 590 (60.3) | 02:43 (01:41–03:53) [00:09–08:09] |
| ≥46 years | 550 (20.9) | 02:33 (01:32–03:40) [00:06–08:25] | 316 (19.1) | 02:34 (01:34–03:43) [00:16–08:25] | 234 (23.9) | 02:32 (01:26–03:36) [00:06–07:47] |
| **Carrying a baby or very young child** | | | | | | |
| No | 2,271 (86.5) | 02:32 (01:34–03:40) [00:06–10:01] | 1,465 (88.6) | 02:29 (01:33–03:36) [00:09–10:01] | 806 (82.8) | 02:39 (01:36–03:48) [00:06–08:09] |
| Yes | 344 (13.1) | 03:04 (01:51–04:07) [00:18–08:21] | 187 (11.3) | 03:09 (01:59–04:22) [00:29–08:21] | 157 (16.1) | 02:55 (01:42–03:58) [00:18–07:15] |
| NR | 12 (0.5) | 03:10 (01:47–04:02) [00:49–06:42] | 4 (0.1) | 01:47 (01:47–01:47) [01:47–01:47] | 11 (1.1) | 03:20 (01:58–04:02) [00:49–06:42] |
| **Attending with ≥1 other person** | | | | | | |
| No | 1,983 (75.3) | 02:31 (01:33–03:39) [00:06–08:53] | 1,279 (77.2) | 02:28 (01:32–03:35) [00:10–08:53] | 704 (72.0) | 02:38 (01:37–03:44) [00:06–08:09] |
| Yes | 651 (24.7) | 02:53 (01:44–03:57) [00:09–10:01] | 377 (22.8) | 02:50 (01:46–03:53) [00:09–10:01] | 274 (28.0) | 02:57 (01:36–03:58) [00:10–08:00] |
| **Time of arrival** | | | | | | |
| 0700–0759 | 737 (28.0)* | 03:08 (02:06–04:19) [00:23–09:44] | 443 (26.7)* | 03:09 (02:05–04:23) [00:30–09:44] | 295 (30.1)* | 03:07 (02:06–04:14) [00:23–08:09] |
| 0800–0859 | 582 (22.1)* | 02:40 (01:41–04:03) [00:10–10:01] | 358 (21.6)* | 02:34 (01:43–03:56) [00:12–10:01] | 224 (22.9)* | 02:49 (01:36–04:13) [00:10–07:28] |
| 0900–0959 | 494 (18.7)* | 02:38 (01:28–03:42) [00:09–07:18] | 299 (18.0)* | 02:41 (01:35–03:44) [00:09–07:18] | 195 (19.9)* | 02:32 (01:19–03:35) [00:09–06:49] |
| 1000–1059 | 320 (12.2)* | 02:38 (01:50–03:28) [00:13–07:30] | 195 (11.8)* | 02:36 (01:43–03:27) [00:13–07:30] | 126 (12.8)* | 02:41 (01:52–03:30) [00:30–05:51] |
| 1100–1159 | 234 (8.9)* | 02:06 (01:19–02:57) [00:06–07:08] | 141 (8.5)* | 02:06 (01:16–02:59) [00:12–07:08] | 93 (9.5)* | 02:06 (01:25–02:57) [00:06–05:23] |
| 1200–1259 | 152 (5.8)* | 01:30 (00:59–02:30) [00:13–06:04] | 116 (7.0)* | 01:34 (01:05–02:36) [00:14–06:04] | 36 (3.7)* | 01:18 (00:53–02:04) [00:13–04:12] |
| ≥1300 | 115 (4.4)* | 01:46 (01:02–02:28) [00:10–06:03] | 105 (6.3)* | 01:47 (01:03–02:27) [00:10–05:59] | 10 (1.0)* | 01:43 (00:55–02:42) [00:10–06:03] |

| **Demographic** | **Overall (n = 2,634)** | | **KwaZulu-Natal (n = 1,656)** | | **Western Cape (n = 978)** | |
|---|---|---|---|---|---|---|
| | n (%) | Median (IQR) [range], HH:MM | n (%) | Median (IQR) [range] , HH:MM | n (%) | Median (IQR) [range], HH:MM |
| **Reported main reason for visit** | | | | | | |
| Acute care/HIV care | 1,526 (57.9) | 02:37 (01:39–03:45) [00:09–10:01] | 1,008 (60.9) | 02:30 (01:36–03:38) [00:09–10:01] | 518 (53.0) | 02:50 (01:48–03:57) [00:10–08:09] |
| Tuberculosis | 145 (5.5) | 02:00 (01:08–02:55) [00:09–07:36] | 78 (4.7) | 01:48 (01:04–02:43) [00:11–07:36] | 67 (6.9) | 02:12 (01:12–03:09) [00:09–06:37] |
| NCDs | 157 (6.0) | 02:34 (01:31–03:45) [00:10–07:40] | 63 (3.8) | 02:28 (01:27–03:45) [00:19–07:40] | 94 (9.6) | 02:39 (01:33–03:48) [00:10–06:57] |
| Mother & child | 297 (11.3) | 02:47 (01:41–03:43) [00:22–08:19] | 214 (12.9) | 02:57 (01:51–03:47) [00:22–08:19] | 83 (8.5) | 02:23 (01:21–03:27) [00:39–06:50] |
| Ante/post-natal | 66 (2.5) | 02:33 (01:32–03:25) [00:12–05:57] | 42 (2.5) | 02:30 (01:29–03:07) [00:12–05:57] | 24 (2.5) | 02:43 (01:40–03:30) [00:28–05:28] |
| Accompanying | 360 (13.7) | 02:45 (01:34–03:58) [00:06–09:28] | 202 (12.2) | 02:42 (01:36–03:58) [00:12–09:28] | 158 (16.2) | 02:51 (01:32–03:57) [00:06–06:55] |
| Proxy | 79 (3.0) | 02:18 (01:22–03:28) [00:11–08:19] | 47 (2.8) | 02:41 (01:33–03:49) [00:11–08:19] | 32 (3.3) | 01:59 (00:58–02:53) [00:23–06:49] |
| NR | 4 (0.2) | 03:50 (03:12–04:39) [01:24–05:57] | 2 (0.1) | 03:58 (03:14–04:40) [03:05–04:45] | 2 (0.2) | 03:50 (03:01–04:23) [01:24–05:57] |

*Mean of 20 imputations

HH: hours; IQR: interquartile range; MM: minutes; NCD: non-communicable disease; NR: not recorded;

#### 3.3. Proportion of time spent indoors vs outdoors

**Supplementary table 7. Proportion of captured time at clinic spent indoors vs. outdoors (n = 2,190 individuals with total time captured of five minutes or longer; n = 10 visits to 9 clinics)**

| Clinic code | ≥1 outdoor waiting area? | Individuals captured, n | Time captured per individual (minutes), median (IQR) [range] | Proportion of time spent indoors (%), median (IQR) [range] | Proportion of captured time spent outdoors (%), median (IQR) [range] | Proportion of captured time spent in unknown location (%), median (IQR) [range] |
|---|---|---|---|---|---|---|
| **All clinics** | | **2,190** | **120.8 (58.3–205.9) [5.1–404.3]** | **95.6 (45.6–100) [0–100]** | **2.5 (0–35.3) [0–100]** | **0 (0–0) [0–100]** |
| Clinics with ≥1 outdoor waiting area | | 1,362 | 126.6 (62.0–126.6) [5.1–404.3] | 73.7 (13.6–97.8) [0–100] | 13.7 (1.4–74.5) [0–100] | 0 (0–0) [0–100] |
| Clinics without an outdoor waiting area | | 828 | 115.2 (51.2–193.7) [5.7–381.6] | 100 (97.1–100) [0–100] | 0 (0–1.4) [0–100] | 0 (0–0) [0–100] |
| **Individual clinics** | | | | | | |
| KZN1 (visit 1) | Yes | 211 | 109.5 (53.1–199.8) [6.5–378.5] | 91.2 (0–99.3) [0–100] | 4.6 (0.5–100) [0–100] | 0 (0–0) [0–100] |
| KZN1 (visit 2) | Yes | 380 | 163.6 (82.8–291.3) [5.1–404.3] | 26.1 (0.8–96.6) [0–100] | 47.2 (2.0–98.7) [0–100] | 0 (0–0) [0–100] |
| KZN2 | Yes | 167 | 161.6 (67.8–217.5) [5.8–352.9] | 56.7 (32.5–82.4) [0–99.4] | 36.2 (9.8–65.0) [0–100] | 0 (0–1.0) [0–97.7] |
| KZN4 | Yes | 262 | 106.7 (58.5–146.9) [5.6–333.1] | 85.6 (50.4–100) [0–100] | 9.4 (0–35.4) [0–99.2] | 0 (0–0) [0–100] |
| KZN5 | Yes | 342 | 132.8 (58.4–203.5) [6.8–370.9] | 86.2 (19.2–97.2) [0–100] | 5.2 (1.7–40.8) [0–100] | 0 (0–3.4) [0–99.7] |
| KZN6 | No | 209 | 85.5 (47.3–132.0) [5.8–323.1] | 97.9 (93.8–99.0) [0–100] | 1.8 (0.8–5.4) [0–100] | 0 (0–0) [0–66.7] |
| WC1 | No | 307 | 145.9 (63.0–230.3) [6.0–381.6] | 100 (99.7–100) [0–100] | 0 (0–0) [0–100] | 0 (0–0) [0–100] |
| WC2 | No | 64 | 124.4 (63.8–193.7) [14.0–363.6] | 98.3 (88.0–99.5) [2.6–99.9] | 0.8 (0.4–3.4) [0–37.6] | 0 (0–1.1) [0–94.3] |
| WC3 | No | 111 | 154.6 (60.3–210.1) [5.8–323.5] | 100 (99.2–100) [33.7–100] | 0 (0–0.6) [0–56.4] | 0 (0–0) [0–66.3] |
| WC6 | No | 137 | 71.1 (24.2–197.2) [5.7–361.1] | 100 (100–100) [22.7–100] | 0 (0–0) [0–77.3] | 0 (0–0) [0–0] |

IQR: interquartile range; KZN: KwaZulu-Natal; WC: Western Cape

**Supplementary table 8. Proportion of captured time at clinic spent indoors vs. outdoors, by self-reported reason for attendance (n = 2,190 individuals with total time captured of five minutes or longer; n = 10 visits to 9 clinics)**

| Clinic code, reason for visit | ≥1 outdoor waiting area? | Individuals captured, n | Time captured per individual (minutes), median (IQR) [range] | Proportion of time spent indoors (%), median (IQR) [range] | Proportion of captured time spent outdoors (%), median (IQR) [range] | Proportion of captured time spent in unknown location (%), median (IQR) [range] |
|---|---|---|---|---|---|---|
| **KZN1 (visit 1)** | **Yes** | **211** | **109.5 (53.1–199.8) [6.5–378.5]** | **91.2 (0–99.3) [0–100]** | **4.6 (0.5–100) [0–100]** | **0 (0–0) [0–100]** |
| Acute care | | 89 | 106.7 (56.4–196.7) [7.8–367.5] | 96.7 (59.7–99.6) [0–100] | 2.9 (0.2–40.3) [0–100] | 0 (0–0) [0–100] |
| HIV/ART | | 38 | 91.2 (42.1–151.9) [7.9–344.9] | 0 (0–7.4) [0–100] | 100 (92.6–100) [0–100] | 0 (0–0) [0–81.4] |
| Other chronic | | 10 | 35.1 (18.5–87.0) [16.0–256.7] | 0 (0–0) [0–100] | 100 (100–100) [0–100] | 0 (0–0) [0–0] |
| TB | | 2 | 46.6 (21.9–71.4) [21.9–71.4] | 0 (0–0) [0–0] | 100 (100–100) [100–100] | 0 (0–0) [0–0] |
| Accompanying a child | | 22 | 208.7 (153.3–295.4) [54.0–378.5] | 98.4 (80.3–99.8) [3.4–100] | 1.6 (0.2–19.7) [0–96.6] | 0 (0–0) [0–0] |
| Accompanying an adult | | 15 | 66.1 (43.1–120.8) [15.9–199.8] | 95.3 (0–98.3) [0–99.4] | 2.0 (0.8–100) [0–100] | 0 (0–0) [0–85.7] |
| Attending for someone else | | 9 | 64.4 (36.2–103.5) [6.5–368.6] | 0 (0–99.3) [0–99.8] | 100 (0.7–100) [0.2–100] | 0 (0–0) [0–0] |
| Mother & child | | 26 | 191.0 (63.2–289.6) [33.0–377.5] | 98.1 (66.1–100) [15.5–100] | 0.9 (0–2.9) [0–66.1] | 0 (0–0) [0–84.5] |
| **KZN1 (visit 2)** | **Yes** | **380** | **163.6 (82.8–291.3) [5.1–404.3]** | **26.1 (0.8–96.6) [0–100]** | **47.2 (2.0–98.7) [0–100]** | **0 (0–0) [0–100]** |
| Acute care | | 118 | 138.8 (80.3–277.6) [5.3–403.4] | 89.8 (18.9–98.3) [0–100] | 7.2 (1.0–50.1) [0–100] | 0 (0–0) [0–100] |
| HIV/ART | | 125 | 144.6 (64.0–294.8) [6.9–404.3] | 1.3 (0–6.4) [0–96.4] | 98.6 (92.8–100) [0–100] | 0 (0–0) [0–10.4] |
| Other chronic | | 18 | 206.0 (103.8–264.9) [10.3–397.9] | 0.1 (0–1.6) [0–10.4] | 99.9 (98.4–100) [89.6–100] | 0 (0–0) [0–0] |
| TB | | 3 | 179.3 (49.2–391.7) [49.2–391.7] | 35.8 (0.3–42.8) [0.3–42.8] | 64.2 (57.2–99.7) [57.2–99.7] | 0 (0–0) [0–0] |
| Accompanying a child | | 43 | 257.1 (165.3–358.6) [14.3–402.7] | 97.5 (81.4–99.3) [0–100] | 2.0 (0.5–6.6) [0–100] | 0 (0–0.3) [0–88.7] |
| Accompanying an adult | | 18 | 102.3 (60–268.3) [5.8–393.7] | 90.4 (4.7–98.0) [0–100] | 5.5 (1.4–53.1) [0–100] | 0 (0–0) [0–100] |
| Attending for someone else | | 11 | 150.4 (85.1–344.1) [37.1–392.7] | 8.2 (0–35.7) [0–95.1] | 91.8 (64.3–100) [4.1–100] | 0 (0–0) [0–0.9] |
| Mother & child | | 42 | 153.6 (84.2–277.7) [5.1–394.3] | 96.7 (68.6–99.4) [0–100] | 1.0 (0.3–5.7) [0–51.8] | 0 (0–1.1) [0–100] |
| Not recorded | | 2 | 223.1 (177.9–268.3) [177.9–268.3] | 51.1 (2.4–99.7) [2.4–99.7] | 48.9 (0.3–97.6) [0.3–97.6] | 0 (0–0) [0–0] |
| **KZN2** | **Yes** | **167** | **161.6 (67.8–217.5) [5.8–352.9]** | **56.7 (32.5–82.4) [0–99.4]** | **36.2 (9.8–65.0) [0–100]** | **0 (0–1.0) [0–97.7]** |
| Acute care | | 43 | 163.6 (79.0–210.6) [8.4–287.2] | 54.6 (30.9–79.4) [0–98.5] | 39.9 (15.1–63.1) [1.5–100] | 0 (0–1.0) [0–97.7] |

| Clinic code, reason for visit | ≥1 outdoor waiting area? | Individuals captured, n | Time captured per individual (minutes), median (IQR) [range] | Proportion of time spent indoors (%), median (IQR) [range] | Proportion of captured time spent outdoors (%), median (IQR) [range] | Proportion of captured time spent in unknown location (%), median (IQR) [range] |
|---|---|---|---|---|---|---|
| HIV/ART | | 56 | 128.0 (54.1–199.0) [7.4–325.1] | 41.4 (31.5–70.6) [0–97.5] | 55.2 (23.9–67.8) [0–100] | 0 (0–0.5) [0–60.3] |
| Other chronic | | 7 | 161.6 (23.6–211.3) [16.3–225.4] | 51.8 (20.9–70.7) [0–91.3] | 48.2 (29.3–73.1) [8.7–100] | 0 (0–0.4) [0–6.0] |
| TB | | 1 | 5.8 (5.8–5.8) [5.8–5.8] | 0.3 (0.3–0.3) [0.3–0.3] | 99.7 (99.7–99.7) [99.7–99.7] | 0 (0–0) [0–0] |
| Accompanying a child | | 11 | 224.3 (40–268.0) [5.9–287.9] | 74.6 (23.8–92.7) [0.4–93.2] | 17.7 (6.8–45.2) [2.6–77.7] | 0 (0–0.7) [0–97.0] |
| Accompanying an adult | | 11 | 64.8 (53.2–165.5) [13.8–320] | 47.8 (0–77.3) [0–98.6] | 26.1 (4.5–95.6) [1.4–100] | 0 (0–4.4) [0–48.8] |
| Attending for someone else | | 5 | 65.2 (51.4–77.3) [7.7–234.1] | 93.3 (52.3–93.5) [42.1–98.5] | 5.7 (1.6–35.1) [1.5–57.4] | 1.1 (0.5–4.9) [0–12.6] |
| Mother & child | | 33 | 202.5 (162.5–234.9) [58.9–352.9] | 77.2 (62.3–94.1) [30.1–99.4] | 14.3 (3.4–35.6) [0.4–69.9] | 0.2 (0–1.3) [0–45.1] |
| **KZN4** | **Yes** | **262** | **106.7 (58.5–146.9) [5.6–333.1]** | **85.6 (50.4–100) [0–100]** | **9.4 (0–35.4) [0–99.2]** | **0 (0–0) [0–100]** |
| Acute care | | 24 | 56.6 (29.2–146.9) [5.6–227.6] | 100 (76.6–100) [32.8–100] | 0 (0–21.1) [0–67.2] | 0 (0–0) [0–17.7] |
| HIV/ART | | 227 | 111.2 (61.2–147.0) [6.4–333.1] | 83.7 (49.9–100) [0–100] | 10.9 (0–35.8) [0–99.2] | 0 (0–0) [0–100] |
| TB | | 5 | 62.8 (29.3–63.4) [28.6–142.3] | 100 (100–100) [96.1–100] | 0 (0–0) [0–3.9] | 0 (0–0) [0–0] |
| Accompanying a child | | 2 | 130.6 (47.4–213.7) [47.4–213.7] | 71.5 (43.1–100) [43.1–100] | 28.5 (0–56.9) [0–56.9] | 0 (0–0) [0–0] |
| Accompanying an adult | | 2 | 92.4 (38.9–145.8) [38.9–145.8] | 67.4 (34.8–100) [34.8–100] | 32.6 (0–65.2) [0–65.2] | 0 (0–0) [0–0] |
| Mother & child | | 2 | 92.9 (47.4–138.4) [47.4–138.4] | 72.0 (44.1–100) [44.1–100] | 23.8 (0–47.7) [0–47.7] | 4.1 (0–8.3) [0–8.3] |
| **KZN5** | **Yes** | **342** | **132.8 (58.4–203.5) [6.8–370.9]** | **86.2 (19.2–97.2) [0–100]** | **5.2 (1.7–40.8) [0–100]** | **0 (0–3.4) [0–99.7]** |
| Acute care | | 99 | 104.2 (58.1–185.1) [6.8–349.7] | 89.0 (16.3–97.8) [0–99.8] | 4.6 (1.3–40.8) [0–100] | 0 (0–3.6) [0–99.7] |
| HIV/ART | | 112 | 156.3 (86.5–202.1) [10.8–370.9] | 91.7 (64.9–97.0) [0–100] | 4.6 (1.8–13.9) [0–100] | 0 (0–3.5) [0–96.9] |
| Other chronic | | 11 | 55.0 (29.6–130.9) [9.3–336.0] | 95.6 (81.5–96.9) [5.1–98.3] | 4.4 (2.7–16.1) [1.7–94.9] | 0 (0–0) [0–26.5] |
| TB | | 20 | 124.2 (74.4–162.5) [11.6–256.5] | 46.0 (18.1–91.8) [0–99.2] | 49.8 (8.2–81.9) [0.8–100] | 0 (0–0) [0–8.3] |
| Accompanying a child | | 11 | 176.0 (90.5–226.1) [12.6–280.7] | 31.5 (0–87.2) [0–99.6] | 37.1 (10.9–100) [0.4–100] | 0 (0–3.4) [0–61.6] |
| Accompanying an adult | | 18 | 89.2 (54.2–177.2) [16.0–214.8] | 65.4 (4.8–98.4) [0.5–99.0] | 2.4 (1.1–43.6) [0–96.9] | 0 (0–37.0) [0–98.2] |
| Attending for someone else | | 10 | 163.0 (41.5–193.9) [13.3–310.3] | 88.2 (32.3–96.2) [0–99.6] | 6.8 (3.3–34.1) [0.4–100] | 0 (0–0) [0–94.0] |
| Mother & child | | 61 | 150.1 (57.2–226.8) [7.1–321.7] | 37.5 (1.7–97.8) [0–99.8] | 7.2 (1.2–72.5) [0–100] | 0 (0–24.3) [0–98.1] |

| Clinic code, reason for visit | ≥1 outdoor waiting area? | Individuals captured, n | Time captured per individual (minutes), median (IQR) [range] | Proportion of time spent indoors (%), median (IQR) [range] | Proportion of captured time spent outdoors (%), median (IQR) [range] | Proportion of captured time spent in unknown location (%), median (IQR) [range] |
|---|---|---|---|---|---|---|
| **KZN6** | **No** | **209** | **85.5 (47.3–132.0) [5.8–323.1]** | **97.9 (93.8–99.0) [0–100]** | **1.8 (0.8–5.4) [0–100]** | **0 (0–0) [0–66.7]** |
| Acute care | | 86 | 66.3 (33.9–125.1) [5.8–323.1] | 96.8 (93.1–98.8) [0–100] | 1.9 (1.0–6.7) [0–100] | 0 (0–0) [0–66.7] |
| HIV/ART | | 30 | 104.9 (35.1–156.9) [13.3–248.1] | 98.5 (95.0–99.1) [34.0–100] | 1.5 (0.7–4.9) [0–66.0] | 0 (0–0) [0–4.1] |
| Other chronic | | 11 | 85.5 (25.2–113.6) [12.3–144.4] | 97.5 (95.0–99.3) [87.4–99.6] | 2.5 (0.7–5.0) [0.4–12.6] | 0 (0–0) [0–0] |
| TB | | 7 | 118.2 (97.0–138.9) [63.9–163.8] | 99.4 (98.4–100) [98.3–100] | 0.6 (0–1.6) [0–1.7] | 0 (0–0) [0–0] |
| Accompanying a child | | 15 | 113.7 (50.4–165.2) [11.5–261.1] | 98.4 (95.7–99.3) [86.4–99.7] | 1.6 (0.7–3.9) [0.3–13.6] | 0 (0–0) [0–4.4] |
| Accompanying an adult | | 12 | 49.2 (19.1–88.0) [7.6–266.4] | 83.1 (58.0–92.9) [17.3–100] | 16.9 (7.1–42.0) [0–82.7] | 0 (0–0) [0–4.6] |
| Attending for someone else | | 4 | 80.8 (47.7–105.2) [27.5–116.7] | 98.3 (95.5–98.6) [92.8–98.8] | 1.7 (1.4–4.5) [1.2–7.2] | 0 (0–0) [0–0] |
| Mother & child | | 44 | 115.1 (61.1–165.9) [7.1–216.7] | 98.1 (95.3–99.2) [61.0–100] | 1.2 (0.6–3.1) [0–39.0] | 0 (0–0) [0–27.1] |
| **WC1** | **No** | **307** | **145.9 (63.0–230.3) [6.0–381.6]** | **100 (99.7–100) [0–100]** | **0 (0–0) [0–100]** | **0 (0–0) [0–100]** |
| Acute care/HIV* | | 172 | 133.1 (62.7–215.8) [6.2–381.6] | 100 (99.3–100) [0–100] | 0 (0–0) [0–100] | 0 (0–0) [0–100] |
| Other chronic | | 52 | 151.4 (58.8–214.3) [8.0–363.9] | 100 (100–100) [0–100] | 0 (0–0) [0–56.4] | 0 (0–0) [0–97.7] |
| TB | | 7 | 213.8 (135.3–258.0) [74.1–325.7] | 100 (100–100) [96.3–100] | 0 (0–0) [0–3.7] | 0 (0–0) [0–0] |
| Accompanying a child | | 18 | 203.4 (55.5–246.1) [33.7–306.6] | 100 (100–100) [98.6–100] | 0 (0–0) [0–1.4] | 0 (0–0) [0–0.8] |
| Accompanying an adult | | 10 | 63.9 (38.3–190.3) [6.0–336.3] | 100 (99.4–100) [93.6–100] | 0 (0–0) [0–0.8] | 0 (0–0) [0–6.4] |
| Attending for someone else | | 17 | 138.1 (31.1–221.0) [8.7–291.4] | 100 (100–100) [91.1–100] | 0 (0–0) [0–8.9] | 0 (0–0) [0–0.6] |
| Mother & child | | 29 | 184.7 (106.9–242.7) [35.2–308.0] | 100 (100–100) [93.5–100] | 0 (0–0) [0–6.5] | 0 (0–0) [0–2.6] |
| Not recorded | | 2 | 233.0 (144.4–321.5) [144.4–321.5] | 99.1 (98.2–100) [98.2–100] | 0.9 (0–1.8) [0–1.8] | 0 (0–0) [0–0] |
| **WC2** | **No** | **64** | **124.4 (63.8–193.7) [14.0–363.6]** | **98.3 (88.0–99.5) [2.6–99.9]** | **0.8 (0.4–3.4) [0–37.6]** | **0 (0–1.1) [0–94.3]** |
| Acute care/HIV* | | 21 | 144.0 (89.1–208.5) [14.0–363.6] | 98.7 (90.5–99.6) [2.6–99.9] | 0.6 (0.4–3.0) [0.1–18.0] | 0 (0–2.0) [0–94.3] |
| TB | | 3 | 118.8 (48.9–153.8) [48.9–153.8] | 98.8 (97.6–99.6) [97.6–99.6] | 1.2 (0.4–2.4) [0.4–2.4] | 0 (0–0) [0–0] |
| Accompanying a child | | 12 | 74.6 (54.8–176.3) [52.2–264.6] | 98.3 (81.9–99.3) [30.5–99.9] | 1.7 (0.7–7.2) [0.1–27.1] | 0 (0–0.3) [0–48.7] |
| Accompanying an adult | | 1 | 58.8 (58.8–58.8) [58.8–58.8] | 81.1 (81.1–81.1) [81.1–81.1] | 0.8 (0.8–0.8) [0.8–0.8] | 18.1 (18.1–18.1) [18.1–18.1] |

| Clinic code, reason for visit | ≥1 outdoor waiting area? | Individuals captured, n | Time captured per individual (minutes), median (IQR) [range] | Proportion of time spent indoors (%), median (IQR) [range] | Proportion of captured time spent outdoors (%), median (IQR) [range] | Proportion of captured time spent in unknown location (%), median (IQR) [range] |
|---|---|---|---|---|---|---|
| Mother & child | | 27 | 123.9 (66.3–193.8) [45.4–302.8] | 98.1 (86.9–99.5) [43.3–99.9] | 0.7 (0.3–6.7) [0–37.6] | 0 (0–0.6) [0–56.6] |
| **WC3** | **No** | **111** | **154.6 (60.3–210.1) [5.8–323.5]** | **100 (99.2–100) [33.7–100]** | **0 (0–0.6) [0–56.4]** | **0 (0–0) [0–66.3]** |
| Acute care/HIV* | | 43 | 172.5 (117.1–200.8) [15.3–323.4] | 100 (99.3–100) [33.7–100] | 0 (0–0) [0–30.5] | 0 (0–0) [0–66.3] |
| TB | | 15 | 32.2 (9.3–46.9) [5.8–250.1] | 100 (99.1–100) [43.6–100] | 0 (0–0) [0–56.4] | 0 (0–0) [0–10.8] |
| Accompanying a child | | 22 | 197.7 (103.8–272.4) [31.6–323.5] | 100 (98.1–100) [93.8–100] | 0 (0–1.9) [0–6.2] | 0 (0–0) [0–2.5] |
| Accompanying an adult | | 4 | 160.2 (69.2–212.6) [5.8–237.4] | 83.0 (55.0–100) [44.0–100] | 16.9 (0–44.9) [0–56.0] | 0 (0–0.2) [0–0.3] |
| Attending for someone else | | 2 | 40.6 (34.0–47.1) [34.0–47.1] | 91.3 (82.7–100) [82.7–100] | 8.7 (0–17.3) [0–17.3] | 0 (0–0) [0–0] |
| Mother & child | | 25 | 167.7 (80.4–201.4) [6.6–284.5] | 100 (100–100) [86.1–100] | 0 (0–0) [0–13.9] | 0 (0–0) [0–0] |
| **WC6** | **No** | **137** | **71.1 (24.2–197.2) [5.7–361.1]** | **100 (100–100) [22.7–100]** | **0 (0–0) [0–77.3]** | **0 (0–0) [0–0]** |
| Acute care/HIV* | | 76 | 121.0 (37.3–246.5) [5.7–361.1] | 100 (100–100) [22.7–100] | 0 (0–0) [0–77.3] | 0 (0–0) [0–0] |
| Other chronic | | 10 | 97.0 (44.5–168.0) [14.8–239.0] | 100 (100–100) [100–100] | 0 (0–0) [0–0] | 0 (0–0) [0–0] |
| TB | | 12 | 23.7 (13.9–50.4) [9.0–225.6] | 100 (100–100) [99.9–100] | 0 (0–0) [0–0.1] | 0 (0–0) [0–0] |
| Accompanying a child | | 13 | 168.9 (71.9–194.8) [9.6–323.9] | 100 (100–100) [97.1–100] | 0 (0–0) [0–2.9] | 0 (0–0) [0–0] |
| Accompanying an adult | | 11 | 58.1 (20.2–112.3) [14.5–137.4] | 100 (100–100) [92.6–100] | 0 (0–0) [0–7.4] | 0 (0–0) [0–0] |
| Attending for someone else | | 9 | 32.1 (22.8–58.3) [6.3–61.5] | 100 (97.3–100) [82.8–100] | 0 (0–2.7) [0–17.2] | 0 (0–0) [0–0] |
| Mother & child | | 6 | 81.3 (20.6–92.9) [17.0–112.0] | 100 (100–100) [97.6–100] | 0 (0–0) [0–2.4] | 0 (0–0) [0–0] |

*Acute care and HIV combined for Western Cape clinics due to error in data collection (see Methods in main article for details)
ART: antiretroviral therapy; IQR: interquartile range; KZN: KwaZulu-Natal; TB: tuberculosis; WC: Western Cape

#### 3.4. Occupancy density

**Supplementary table 9. Occupancy density by room area and room volume in three indoor spaces in each of clinics KZN2, KZN6, and WC1 from 0800–1345***

| Clinic | Space | Designation | Area, $m^2$ | Mean height, m† | Maximum height, m | Volume, $m^3$ | Occupancy (n), median (IQR) [range] | Occupancy density by room area (persons/$m^2$), median (IQR) [range] | Occupancy density by room volume (persons /$m^3$), median (IQR) [range] | Comments |
|---|---|---|---|---|---|---|---|---|---|---|
| **KZN6** | A | Pre-registration/pre-vitals waiting area | 65.6 | 2.5 | 2.5 | 162.8 | 41 (35–45) [23–57] | 0.63 (0.53–0.69) [0.35–0.87] | 0.25 (0.21–0.28) [0.14–0.35] | All spaces have relatively low, flat ceilings |
| | B | Pre-consultation waiting area | 19.4 | 2.5 | 2.5 | 48.2 | 19 (11–27) [0–32] | 0.98 (0.57–1.39) [0–1.65] | 0.39 (0.23–0.56) [0–0.66] | |
| | C | Pre-consultation waiting area | 15.9 | 2.5 | 2.5 | 25.6 | 26 (21–28) [11–34] | 1.63 (1.32–1.76) [0.69–2.14] | 1.02 (0.82–1.09) [0.43–1.33] | |
| **KZN2** | D | Pre-registration waiting area | 49.7 | 2.7 | 2.7 | 136.4 | 30 (19–34) [7–51] | 0.60 (0.38–0.68) [0.14–1.03] | 0.22 (0.14–0.25) [0.05–0.37] | All spaces have relatively low, flat ceilings |
| | E | Corridor used as pre-consultation waiting area | 20.4 | 2.6 | 2.6 | 42.7 | 11 (9–13) [0–19] | 0.54 (0.44–0.64) [0–0.93] | 0.21 (0.17–0.24) [0–0.36] | |
| | F | Combined pre-vitals waiting area, vitals administration area, and registration area | 16.3 | 2.6 | 2.6 | 54.4 | 14 (9–19) [0–29] | 0.86 (0.55–1.16) [0–1.78] | 0.33 (0.21–0.45) [0–0.68] | |
| **WC1** | G | Pre-registration waiting area | 129.5 | 3.5 | 4.2 | 368.2 | 52 (45–56) [29–71] | 0.40 (0.35–0.43) [0.22–0.55] | 0.13 (0.10–0.15) [0–0.19] | All spaces have relatively high, sloping ceilings |
| | H | Pre-vitals waiting area | 53.2 | 5.3 | 5.9 | 272.8 | 29 (21–34) [5–43] | 0.55 (0.39–0.64) [0.09–0.81] | 0.10 (0.07–0.12) [0–0.16] | |
| | I | Pre-consultation waiting area | 37.7 | 4.2 | 5.9 | 169.1 | 23 (18–25) [8–34] | 0.61 (0.48–0.66) [0.21–0.90] | 0.14 (0.09–0.15) [0–0.20] | |

*Data available only from 0830 for clinic KZN6
†Some spaces had sloping ceilings and/or areas with lower flat ceilings
IQR: interquartile range

#### Appendix 4. Additional discussion

##### Supplementary table 10. Summary of previous studies estimating waiting times in South African PHC clinics

| First author, year published | Location | Number of facilities | Facility type | Dates of data collection | Data collection methods | Key findings |
|---|---|---|---|---|---|---|
| Stime, 2018[17] | Durban, KZN | 1 | HIV/TB/STI outpatient centre | 2016 | Time and motion (direct observation) | Median time in clinic 01:48 (range 00:28–04:35) for STI care (n = 39) and 02:46 (range 01:25–03:50) for those attending for HIV care (n = 27). HIV 'fast track' slightly reduced time (median 02:24 [range 00:14–04:43; n = 28]) and use of point-of-care diagnostics for STIs (instead of syndromic management) increased time spent (median 04:02; n = 9). |
| Egbujie, 2018[18] | KZN | 9 | PHC clinics | 2014–15 | Adapted NDoH waiting time approach | Implemented a range of Ideal Clinic recommendations.<br>Pre-intervention median time in clinic 01:56 (IQR 01:06–02:48); n = 860<br>Post-intervention median time in clinic 02:02 (IQR 01:21–03:24); n = 903<br>Multivariable analysis: longer waiting times associated with earlier arrival time and higher patient/nurse ratio. |
| Daniels, 2017[19] | WC | 22 | PHC clinics | 2007, 2011 | Standard WTSE approach | 2007 survey: median waiting time* 01:16 (IQR 00:37–02:05); median 233 patients attending per day per clinic<br>2011 survey: median waiting time* 00:55 (IQR 00:28–01:54); median 255 patients attending per day per clinic |
| Bachmann, 1997[1] | Khayelitsha, WC | 1 | CHC | 1993 | Paper-based waiting time approach | Median time in clinic: 2.6 (IQR 1.7–3.8) hours and 4.1 (IQR 2.9–4.9) hours for those attending for 'preventive' (n = 368) and 'curative' (n = 416) care, respectively. |

*Only time spent waiting, does not include time spent receiving services
CHC: community health centre; IQR: interquartile range; KZN: KwaZulu-Natal; NDoH: National Department of Health; PHC: primary health care; STI: sexually transmitted infection; TB: tuberculosis; WC: Western Cape; WTSE: Waiting Time and Systems Efficiency

### Appendix 5. Acknowledgments

##### Supplementary table 11. The extended *Umoya omuhle* team, institutions, and roles (listed alphabetically by surname)

| Name | Institution/s | Role |
|---|---|---|
| Siphokazi Adonisi | UCT | Research Assistant |
| Kathy Baisley | LSHTM; AHRI | Co-investigator |
| Peter Beckwith | LSHTM; UCT | Research fellow |
| Fiammetta Bozzani | LSHTM | Co-investigator |
| Amy Burdzik | UCT | Occupational health |
| Adrienne Burrough | LSHTM | Project Manager |
| Nkosingiphile Buthelezi | AHRI | Research Assistant |
| Xolile Buthelezi | AHRI | Diagnostic Lab Manager |
| Ruvimbo Chigwanda | UCT | Administration |
| Christopher Colvin | UCT | Co-investigator |
| PIP CRAs | AHRI | Clinic research Assistants |
| Njabulo Dayi | AHRI | Research Data Manager |
| Arminder Deol | LSHTM | Mathematical modeller |
| Karina Diaconu | QMU | Co-investigator |
| Siphephelo Dlamini | AHRI | Nursing Manager |
| Yutu Dlamini | AHRI | Research Assistant |
| Raveshni Durgiah | AHRI | Grants office |
| Anita Edwards | AHRI | Head: Scientific Support |
| Jennifer Falconer | QMU | Research Assistant |
| Kitty Flynn | QMU | Administrator |
| Patrick Gabela | AHRI | Clinical Research Data Coordinator |
| Dickman Gareta | AHRI | Head: Research Data Management |
| Awethu Gawulekapa | UCT | Research Assistant |
| Harriet Gliddon | AHRI; UCL | Research Assistant |
| Bavashni Govender | UKZN | Administration |
| Indira Govender | LSHTM; AHRI | Co-investigator |
| Alison Grant | LSHTM; AHRI | Principal investigator |
| Meghann Gregg | LSE | Research Fellow |
| Emmerencia Gumede | AHRI | Research Assistant |
| Sashin Harilall | AHRI | Grants office |
| Kobus Herbst | AHRI | Chief Information Officer |
| Tamia Jansen | UCT | Research Assistant |
| Seonaid Kabiah | UCT | Research Assistant |
| Idriss Kallon | UCT | Post-doctoral researcher |
| Aaron Karat | LSHTM | Co-investigator |
| Hannah Keal | AHRI | Communications |
| Suzanne Key | UCT | Occupational health |
| Zama Khanyile | UKZN | Research Assistant |
| Mandla Khoza | AHRI | Clinic Research Assistant |
| Nozi Khumalo | AHRI | Systems Engineer |

| Name | Institution/s | Role |
|---|---|---|
| Zilethile Khumalo | AHRI | Research Assistant |
| Karina Kielmann | QMU | Co-principal investigator |
| Nondumiso Kumalo | AHRI | Clinic Research Assistant |
| Richard Lessells | AHRI | Epidemiologist |
| Nokuthula Lushaba (deceased) | UKZN | Administration |
| Sithembiso Luthuli | AHRI | Research Assistant |
| Sinethemba Mabuyakhulu | AHRI | Clinic Research Assistant |
| Hayley MacGregor | IDS | Co-investigator |
| Nonhlanhla Madlopha | AHRI | Research Assistant |
| Aphiwe Makalima | UCT | Administration |
| Tacha Malaza | AHRI | PIP CRA |
| Sifundesihle Malembe | AHRI | Research Assistant |
| Godfrey Manuel | UCT | Transport |
| Nonhlanhla Maphumulo | UKZN | Administration |
| Precious Mathenjwa | UCT | Research Assistant |
| Sanele Mbuyazi | AHRI | PIP CRA |
| Nicky McCreesh | LSHTM | Co-investigator |
| Claire McLellan | QMU | Administrator |
| Simphiwe Mdluli | AHRI | PIP CRA |
| Thabile Mkhize | AHRI | Transport |
| Duduzile Mkhwanazi | AHRI | Research Assistant |
| Zinhle Mkhwanazi | AHRI | Research Assistant |
| Zodwa Mkhwanazi | AHRI | Research Assistant |
| Anathi Mngxekeza | UCT | Research Assistant |
| Tshwaraganang Modise | AHRI | Research Data |
| Sashen Moodley | AHRI | Microbiology Laboratory Supervisor |
| Samantha Moyo | UCT | Research Assistant |
| Silindile Mthembu | AHRI | Clinic Research Assistant |
| Nozipho Mthethwa | AHRI | Research Assistant |
| Siphesihle Mthethwa | AHRI | Procurement Coordinator |
| Sphiwe Mthethwa | AHRI | Research Assistant |
| Sanele Mthiyane | AHRI | Research Assistant |
| Vanisha Munsamy | AHRI | Grants office |
| Sinead Murphy | UCT | Research Assistant |
| Thomas Murray | AHRI | Research assistant |
| Senzile Myeni | AHRI | PIP CRA |
| Tevania Naidoo | AHRI | Procurement |
| Nompilo Ndlela | AHRI | Research Assistant |
| Zama Ndlela | AHRI | PIP CRA |
| Thandekile Nene | AHRI | Research Assistant |
| Phumla Ngcobo | AHRI | Communications |
| Nzuzo Ntombela | AHRI | Research Data Systems Service Manager |
| Sabelo Ntuli | AHRI | GIS Coordinator |
| Nompumulelo Nyawo | AHRI | Human resources |

| Name | Institution/s | Role |
|---|---|---|
| Phumzile Nywagi | UCT | Research Assistant |
| Stephen Olivier | AHRI | Statistician |
| Justin Parkhurst | LSE | Co-investigator |
| Alex Pym | AHRI | Co-investigator |
| Yolanda Qeja | UCT | Research Assistant |
| Anand Ramnanan (deceased) | AHRI | Procurement |
| Sharmila Rugbeer | UKZN | Administration |
| Janet Seeley | LSHTM | Co-investigator |
| Aruna Sevakram | AHRI | Scientific support |
| Sizwe Sikhakane | AHRI | Transport |
| Zizile Sikhosana | AHRI | Somkhele Laboratory Supervisor |
| Theresa Smit | AHRI | Head: Diagnostic Research |
| Thandeka Smith | UKZN | Research Assistant |
| Naomi Stewart | LSHTM | Communications |
| Alison Swartz | UCT | Co-investigator |
| Amy Thomas | LSHTM | Communications |
| Siphosethu Titise | UCT | Research Assistant |
| Anna Vassall | LSHTM | Co-investigator |
| Marlise Venter | AHRI | Facilities Administrator |
| Anna Voce | UKZN | Co-investigator |
| Richard White | LSHTM | Co-investigator |
| Tom Yates | Imperial | Co-investigator |
| Precious Zulu | AHRI | Administration |
| Gimenne Zwama | QMU | Research Fellow |

AHRI: Africa Health Research Institute; IDS: Institute of Development Studies; LSE: London School of Economics and Political Science; LSHTM: London School of Hygiene & Tropical Medicine; QMU: Queen Margaret University; UCT: University of Cape Town; UKZN: University of KwaZulu-Natal;

### Appendix 6. References

1 Bachmann MO, Barron P. Why wait so long for child care? An analysis of waits, queues and work in a South African urban health centre. *Trop Doct*. 1997;**27**(1):34–8.

2 South Africa National Department of Health. Ideal Clinic Integrated Clinical Services Management Manual. https://www.idealhealthfacility.org.za/docs/Integrated%20Clinical%20Services%20Management%20%20Manual%205th%20June%20FINAL.pdf (accessed 2010 Jul 2)

3 Reagon G, Igumbor E. Strengthening Health Systems through training of Health Care Providers in the conduct of Routine Waiting Time and System Efficiency Surveys. *Stud Health Technol Inform*. 2010;590–4.

4 Singman EL, Haberman CV, Appelbaum J, et al. Electronic Tracking of Patients in an Outpatient Ophthalmology Clinic to Improve Efficient Flow: A Feasibility Analysis and Benchmarking Study. *Qual Manag Health Care*. 2015;**24**(4):190–9.

5 Montanari A, Tian Z, Francu E, et al. Measuring Interaction Proxemics with Wearable Light Tags. *Proc ACM Interact Mob Wearable Ubiquitous Technol*. 2018;**2**(1):1–30.

6 Aharony N, Pan W, Ip C, Khayal I, Pentland A. Social fMRI: Investigating and shaping social mechanisms in the real world. *Pervasive Mob Comput*. 2011;**7**(6):643–59.

7 Aiello AE, Simanek AM, Eisenberg MC, et al. Design and methods of a social network isolation study for reducing respiratory infection transmission: The eX-FLU cluster randomized trial. *Epidemics*. 2016;**15**:38–55.

8 Rodriguez M, Sivic J, Laptev I, Audibert J-Y. Data-driven crowd analysis in videos. In: *2011 International Conference on Computer Vision*. 2011. p. 1235–42.

9 Rodriguez M, Laptev I, Sivic J, Audibert J-Y. Density-aware person detection and tracking in crowds. In: *2011 International Conference on Computer Vision*. 2011. p. 2423–30.

10 Rodriguez M, Ali S, Kanade T. Tracking in unstructured crowded scenes. In: *2009 IEEE 12th International Conference on Computer Vision*. 2009. p. 1389–96.

11 Committee on Traffic Flow Theory and Characteristics, Federal Highway Administration. Traffic Flow Theory. 1975. http://tft.eng.usf.edu/docs.htm (accessed 2021 Apr 12)

12 Armony M, Israelit S, Mandelbaum A, Marmor YN, Tseytlin Y, Yom-Tov GB. On Patient Flow in Hospitals: A Data-Based Queueing-Science Perspective. An Extended Version. *Stoch Syst*. 2015;**5**(1):146–94.

13 Carmen R sa, Van Nieuwenhuyse I. Improving Patient Flow in Emergency Departments with OR Techniques: A Literature Overview. *SSRN Electron J*. 2014;

14 Higginson I. Emergency department crowding. *Emerg Med J*. 2012;**29**(6):437–43.

15 Bean DM, Taylor P, Dobson RJB. A patient flow simulator for healthcare management education. *BMJ Simul Technol Enhanc Learn*. 2019;**5**(1):46–8.

16 McAdams D. Changing the Game of Patient Experience: Insights from Game Theory. 2014. https://faculty.fuqua.duke.edu/~dm121/press/HSAC%20Patient%20Experience%20Final.pdf (accessed 2021 Apr 12)

17 Stime KJ, Garrett N, Sookrajh Y, et al. Clinic flow for STI, HIV, and TB patients in an urban infectious disease clinic offering point-of-care testing services in Durban, South Africa. *BMC Health Serv Res*. 2018;**18**(1):363.

18 Egbujie BA, Grimwood A, Mothibi-Wabafor EC, et al. Impact of 'Ideal Clinic' implementation on patient waiting time in primary healthcare clinics in KwaZulu-Natal Province, South Africa: A before-and-after evaluation. *S Afr Med J*. 2018;**108**(4):311.

19 Daniels J, Zweigenthal V, Reagon G. Assessing the impact of a waiting time survey on reducing waiting times in urban primary care clinicss in Cape Town, South Africa. *J Public Health Afr*. 2017;**8**:23–9.
