## Supplementary material for "Waiting times, patient flow, and occupancy density in South African primary health care clinics: implications for infection prevention and control": STROBE checklist

**STROBE Statement—Checklist of items that should be included in reports of *cross-sectional studies***

| | Item No | Recommendation | Line/page references |
|---|---|---|---|
| **Title and abstract** | 1 | (*a*) Indicate the study's design with a commonly used term in the title or the abstract | - |
| | | (*b*) Provide in the abstract an informative and balanced summary of what was done and what was found | Page 2, lines 7–22 |
| **Introduction** | | | |
| Background/rationale | 2 | Explain the scientific background and rationale for the investigation being reported | Page 4, lines 33–49<br>Page 5, lines 59-68 |
| Objectives | 3 | State specific objectives, including any prespecified hypotheses | Page 5, lines 70–74 |
| **Methods** | | | |
| Study design | 4 | Present key elements of study design early in the paper | Page 5, lines 76–82<br>Appendix 1.1 (page 2 of supplement) |
| Setting | 5 | Describe the setting, locations, and relevant dates, including periods of recruitment, exposure, follow-up, and data collection | Pages 6–7, lines 83–122 |
| Participants | 6 | (*a*) Give the eligibility criteria, and the sources and methods of selection of participants | Page 6, lines 87–89; 103 |
| Variables | 7 | Clearly define all outcomes, exposures, predictors, potential confounders, and effect modifiers. Give diagnostic criteria, if applicable | Pages 8–9, lines 130–170 |
| Data sources/ measurement | 8* | For each variable of interest, give sources of data and details of methods of assessment (measurement). Describe comparability of assessment methods if there is more than one group | Page 6, lines 89–90; 96–102<br>Page 7, lines 105–123 |
| Bias | 9 | Describe any efforts to address potential sources of bias | Page 8, lines 137–144<br>Page 9, lines 156–58; 165–66<br>Page 18, lines 369–377 |
| Study size | 10 | Explain how the study size was arrived at | Page 10, lines 192–194 |
| Quantitative variables | 11 | Explain how quantitative variables were handled in the analyses. If applicable, describe which groupings were chosen and why | Pages 8–9, lines 155–170 |
| Statistical methods | 12 | (*a*) Describe all statistical methods, including those used to control for confounding | Pages 8–9, lines 144–154<br>Appendix 2.2.1 (pages 10–12 of supplement) |
| | | (*b*) Describe any methods used to examine subgroups and interactions | n/a |
| | | (*c*) Explain how missing data were addressed | Page 8, lines 138–142<br>Appendix 2.2.1 (pages 10–12 of supplement) |

| | Item No | Recommendation | Line/page references |
|---|---|---|---|
| | | (*d*) If applicable, describe analytical methods taking account of sampling strategy | n/a |
| | | (*e*) Describe any sensitivity analyses | n/a |
| **Results** | | | |
| Participants | 13* | (a) Report numbers of individuals at each stage of study—eg numbers potentially eligible, examined for eligibility, confirmed eligible, included in the study, completing follow-up, and analysed | Page 10, lines 192–194<br>Page 11, lines 206–209<br>Page 12, line 228; 243<br>Supplementary table 3 (page 10 of supplement)<br>Appendix 3.1 (page 13 of supplement) |
| | | (b) Give reasons for non-participation at each stage | Supplementary table 3 (page 10 of supplement) |
| | | (c) Consider use of a flow diagram | n/a |
| Descriptive data | 14* | (a) Give characteristics of study participants (eg demographic, clinical, social) and information on exposures and potential confounders | Pages 10–11, lines 192–202<br>Table 1 |
| | | (b) Indicate number of participants with missing data for each variable of interest | Table 1<br>Supplementary table 4 (page 12 of supplement) |
| Outcome data | 15* | Report numbers of outcome events or summary measures | Pages 11–12, lines 206–226<br>Table 2<br>Supplementary tables 6, 7, & 8 (pages 15–21 of supplement) |
| Main results | 16 | (*a*) Give unadjusted estimates and, if applicable, confounder-adjusted estimates and their precision (eg, 95% confidence interval). Make clear which confounders were adjusted for and why they were included | Page 11, lines 212–216<br>Table 2 |
| | | (*b*) Report category boundaries when continuous variables were categorized | Table 2 |
| | | (*c*) If relevant, consider translating estimates of relative risk into absolute risk for a meaningful time period | n/a |
| Other analyses | 17 | Report other analyses done—eg analyses of subgroups and interactions, and sensitivity analyses | Pages 12–13, lines 227–260 |
| **Discussion** | | | |
| Key results | 18 | Summarise key results with reference to study objectives | Page 13, lines 262–269 |
| Limitations | 19 | Discuss limitations of the study, taking into account sources of potential bias or imprecision. Discuss both direction and magnitude of any potential bias | Page 18, lines 367–382 |

| | Item No | Recommendation | Line/page references |
|---|---|---|---|
| Interpretation | 20 | Give a cautious overall interpretation of results considering objectives, limitations, multiplicity of analyses, results from similar studies, and other relevant evidence | Pages 14–15, lines 270–303 |
| Generalisability | 21 | Discuss the generalisability (external validity) of the study results | Page 18, lines 377–382 |
| **Other information** | | | |
| Funding | 22 | Give the source of funding and the role of the funders for the present study and, if applicable, for the original study on which the present article is based | Page 20, lines 416–422 |

*Give information separately for exposed and unexposed groups.

**Note:** An Explanation and Elaboration article discusses each checklist item and gives methodological background and published examples of transparent reporting. The STROBE checklist is best used in conjunction with this article (freely available on the Web sites of PLoS Medicine at http://www.plosmedicine.org/, Annals of Internal Medicine at http://www.annals.org/, and Epidemiology at http://www.epidem.com/). Information on the STROBE Initiative is available at www.strobe-statement.org.
